# The impact of antibiotics on the presence of the protozoan anaerobe *Blastocystis* and the surrounding microbiome: a case study

**DOI:** 10.1101/2024.07.25.24310942

**Authors:** Jamie M. Newton, William JS. Edwards, Gary Thompson, Eleni Gentekaki, Anastasios D. Tsaousis

## Abstract

**Background:** *Blastocystis*, the most prevalent eukaryotic gut microbe in humans, has a global distribution. Studies have linked its presence with distinct gut microbiome and metabolome profiles compared to those where the organism is absent. However, the in vivo effect of antibiotics on *Blastocystis* and the surrounding gut microbiome remains understudied. This case study aimed to explore how antibiotic consumption influences the presence of *Blastocystis* and the subsequent changes in the gut microbiome and metabolome of an individual with irritable bowel syndrome (IBS).

**Methods:** Stool samples from an IBS patient, collected at various time points, were tested for *Blastocystis* presence using RT-PCR targeting the *SSUrRNA* gene, followed by sequencing of positive samples. Illumina sequencing determined the gut microbiome composition, while one-dimensional proton NMR spectroscopy analysed the metabolome composition. Statistical analyses were conducted to identify relationships between antibiotic consumption, bacterial diversity, metabolome composition, and *Blastocystis* presence.

**Results:** Antibiotics significantly impacted the gut microbiome, with diversity declining early in the antibiotic course, then recovering later and post-course. *Blastocystis* was detected early, late, and post-course but not mid-course, coinciding with the decline in bacterial diversity. No significant differences were observed between *Blastocystis*-positive and *Blastocystis*-negative samples. However, bacterial composition significantly differed between samples collected before, early, and after the antibiotic course compared to those collected mid-course. Metabolite groups, including short-chain fatty acids, amino acids, and succinate, exhibited changes throughout the antibiotic course, indicating that gut metabolite composition is affected by antibiotic consumption.

**Discussion/Conclusion:** While antibiotics did not significantly impact *Blastocystis* colonisation, they did cause a mid-course decline in microbial diversity and *Blastocystis* presence. The study also revealed significant alterations in important metabolites such as SCFAs and amino acids throughout the antibiotic course, with an altered metabolome observed post-course. This case study underscores the complex interactions between antibiotics, gut microbiota, and metabolites, highlighting the resilience of *Blastocystis* in the gut ecosystem.

## Introduction

The gut microbiome and metabolome are crucial influencers of gastrointestinal (GI) health, and their interactions can be better understood by studying them in tandem [1]. This dual approach is particularly important when investigating GI health differences across various cohorts or when external factors influence the GI tract. For example, a previous metabolomics study using ^1H Nuclear Magnetic Resonance (NMR) spectroscopy revealed significant differences in stool metabolite composition between individuals with diarrhoea and healthy controls and between *Blastocystis* carriers and non-carriers [2]. Understanding these interactions can provide deeper insights into the complex dynamics of gut health and disease.

*Blastocystis* is a eukaryotic microbe that resides in the GI tract and has a global distribution in a broad range of animal hosts [3], [4]. Epidemiological studies and phylogenetic analysis of the small subunit ribosomal RNA (*SSU* rRNA) gene have revealed over 44 different subtypes, twelve of which STs 1-9, 12, 16, 23 have been identified in human stool samples [5], [6], [7], [8], [9]. *Blastocystis* was initially designated as a parasite and linked with IBS and other gastrointestinal disorders [10], [11]; however, more recent studies have indicated a negative correlation between the presence of the organism and gastrointestinal symptoms [8], [9], [12], [13], () muddling its association with disease. *Blastocystis*’ genetic diversity further complicates interpretations, with many studies finding no relationship between inter and intra-subtype diversity and disease [9], [12].

Nonetheless, specific microbial profiles have been associated with the organism. For instance, *Blastocystis* presence is more common in the *Ruminococcaceae* and *Prevotella* enterotypes, rather than *Bacteroides*, and associated with higher richness and diversity, which can be an indicator of good GI health [8], [14], [15]. At the level of subtype, *Blastocystis* ST3 and ST4 have been shown to have an inverse relationship with *Akkermansia* abundance, an indicator of GI health [8]. Whether *Blastocystis* is a gut ecosystem engineer, a simple colonizer or just a passenger is still unknown.

Individuals with irritable bowel syndrome (IBS) have been known to have distinct gut bacterial compositions/profiles to their non-IBS counterparts, making IBS treatment with antibiotics a potential influencing factor [16], [17], [18], [19]. *In vivo* and *in vitro* studies have indicated that the gut microbiomes of individuals colonised with *Blastocystis* show a decline in abundance of genera such as *Bifidobacterium* and *Lactobacillus* [15], [20], [21]. *Bifidobacterium* has a role in immunomodulation and protection of the GI epithelial cells [20], [22], [23]. Therefore, the results of these studies could link *Blastocystis* to dysbiosis-induced GI symptoms and possibly IBS.

Antibiotic administration has been an effective treatment for some IBS cases and other GI conditions, while metronidazole, ciprofloxacin and rifaximin have been effective at decreasing the severity of symptoms in many clinical trials [18], [19], [24], [25], [26]. However, as our understanding of the gut microbiome’s role in gastrointestinal health deepens, the impact of antibiotics on gut microbiota modulation is receiving closer scrutiny. Several studies have detected significant changes in the gut microbiome composition during antibiotic treatment and, occasionally, microbiome recovery after treatment [27], [28], [29].

In this case study, we monitored the metabolome and the bacterial gut microbiome composition of a *Blastocystis*-positive IBS patient during a 14-day course of antibiotics. We also analysed *Blastocystis* presence over this 14-day period and how it is impacted by antibiotic treatment. The subject’s gut microbiome and metabolome composition were also analysed following the termination of the antibiotic course to monitor microbial diversity recovery after *Blastocystis* detection.

## Materials and Method

### Ethics approval

The study was conducted within the guidelines established in IRAS ethics approvals 274985 and 286641, following a review by an ethics committee and applying suggested amendments to comply with ethical standards. The UK National Ethic committees of Health Research Authority (HRA) and Health and Care Research Wales (HCRW) under the umbrella of NHS Health Research Authority gave ethical approval of this work.

### Participant recruitment and sample collection

The study subject, previously diagnosed with IBS, presented to a hospital in the Kent county (South East England) with gastrointestinal symptoms and was put on a 14-day course of antibiotics. These included 500 mg Amoxicillin (a 3^rd^ generation penicillin antibiotic) x 2 and 500 mg Clarithromycin (a 2^nd^ generation macrolide antibiotic). Daily doses of 30 mg Lansoprazole (proton pump inhibitor) x 2 was also prescribed. The subject was in their 40s (41 to 45) with a BMI of 29 and a mixed diet. The subject was provided with faeces catchers (Zymo Research Cat No R1101-1-10) and two types of collection tubes, one containing 5 ml DNA/RNA shield (Zymo Research Cat No R1100-250) and the other containing 5 ml of 50% methanol. Each faecal sample was distributed in the two tubes. Faecal samples were collected just before the commencement of the treatment course, then on D2 then once daily for the remainder of the first week, then once on D8, D10 and D15 (the day after the completion of the course) for the following week. Follow-up samples were then collected at D30 and D3M (3 months after course start). The samples were stored in their respective tubes in DNA/RNA shield or methanol at -80°C.

### DNA extraction

200 mg solid stool stored in DNA/RNA shield or 200 µl liquid stool were added to 200 µl PBS (pH 7.4 RNAase free). The samples were then centrifuged for 10 minutes at 10,000 x g at room temperature (RT). The pellet was then resuspended in the supernatant and the DNA was extracted using the QIAamp PowerFecal Pro DNA Kit (Qiagen; Cat. No: 51804) following the manufacturer’s protocol, and 50 µl DNA was eluted.

### qPCR and *Blastocystis* detection

For *Blastocystis* detection, a 350 bp region of the *SSU* rRNA gene was targeted using a reaction mixture of 2 µl DNA, 500 nM of primer set PPF1 (fwd) (5’-AGTAGTCATACGCTCGTCTCAAA-3’) and R2PP (rvs) (5’-TCTTCGTTACCCGTTACTGC-3’) and 5 µl SYBR green making a full reaction volume of 10 µl. The qPCR was run on a Quantstudio-3 real-time PCR machine with the following program: initial denaturation 95 °C for 5 minutes, then 45 cycles of initial denaturation 95°C for 5 seconds, annealing 68 °C for 10 seconds, extension 72°C 10 seconds then a final extension of 72 °C for15 seconds.

### Sequencing and Subtype annotation

Bi-directional Sanger sequencing using the set of primers for the qPCR reaction was outsourced to and performed by Eurofins (UK). The forward and reverse nucleotide sequences were then assessed and trimmed using SnapGene Viewer Version 6.2.2 (https://www.snapgene.com/snapgene-viewer). The final trimmed consensus sequences were then used as queries to check for contamination using the Basic Local Alignment Search Tool (BLAST) from the National Centre for Biotechnology Information (NCBI) (https://blast.ncbi.nlm.nih.gov/Blast.cgi). Once the identity of the sequence was confirmed as *Blastocystis*, the subtype was assigned using the curated database pubMLST (https://pubmlst.org/organisms/blastocystis-spp).

### 16S rRNA gene amplicon sequencing

Novogene outsourced the high-throughput amplicon sequencing. The protocol used was based on Caporaso et al [30]. [] with some modifications. One ng DNA from extracts was used, fragmented, and then adapted for paired-end sequencing. The DNA was amplified using the primer pair 515F GTGCCAGCMGCCGCGGTAA and 907R CCGTCAATTCCTTTGAGTTT, which amplifies the hypervariable region and then sequenced on the Illumina NovaSeq platform.

The raw reads were classified using the Lotus2 software [[31]]. The parameters and tools used are as follows: Chimera checking/removal was performed using Minimap2 [[32]], and Minimap2 was also used to look for off-target hits containing human DNA ‘contaminated’ reads by BLASTing reads against Genome Reference Consortium Human Build 38.p14. V3-V4 region trimmed reads were then clustered into ASVs (≤ 1 nucleotide dissimilarity) using Divisive Amplicon Denoising Algorithm 2 (DADA2) [[33]], ASVs were taxonomically classified (to species level) using BLAST against the GreenGenes2 (GG2) database [[34]].

GG2 was chosen for its reliability (GG2 is a unified database suitable for whole genome sequencing (WGS) data and 16s data), as well as replicable results.

### Statistical analysis

Statistical analysis and data visualisation were done using the R Studio 4.2.3 package. Relative abundances of each genus were calculated in each sample, and a heatmap was constructed. Diversity index values were calculated using the Phyloseq package. Shannon, Chao1, Simpson and observed taxa values were used. These four values were analysed for statistical differences occurring between the *Blastocystis* positive and *Blastocystis* negative samples, as well as differences in diversity score between the ‘antibiotic positive’ time points (days 4-15) and the ‘antibiotic negative’ time points (day 0, 30 days post antibiotics and 3 months post antibiotics). First, a Shapiro test was used to determine the data distribution to analyse the statistical differences between the sample groups. Normally distributed data was analysed with ANOVA test followed by Tukey HSD test for pairwise comparison. For samples with a non-normal distribution, the Kruskal-Wallis test was used, followed up by the Dunn Test (Bonferroni P-adjust) for pairwise comparisons. The raw diversity index values were also plotted over time. To visualise microbiome composition, compositional plots showing all taxa making up >1% of the total read counts were produced using the Microbiome package. To look for the presence of ‘bio-markers’ of the presence of Blastocystis, Linear Discriminant Effect Size (LEfSe) analysis was done [35]. LEfSe uses a combination of statistical tests to identify taxa whose high/low abundance or presence/absence allows for the best linear discrimination/explanation of the differences observed (changes in taxa) between the 2 groups of samples (*Blastocystis* +ve/-ve). Principle Component Analysis (PCA) was also used to determine differences between the *Blastocystis* +ve/-ve groups based on overall taxa presence and distribution. Samples were plotted based on their Dissimilarity matrix values (Euclidian distances), and principal component analysis was performed. Statistical analysis was then done using PERMANOVA [[34]] to determine if the ‘centrons’ of each group (*Blastocystis* +ve/-ve) differed significantly in location.

### Metabolite extraction

200 mg solid stool stored in methanol or 200 µl liquid stool was resuspended in 4 ml methanol, then 200 mg glass beads were added and vortexed for 30 seconds. The samples were then incubated at RT for 3 minutes, then vortexed for a further 30 seconds. The supernatants were then divided in 4 x 1m aliquots then centrifuged at 10,000 x g at 4°C for 20 mins then lyophilized. The lyophilized desiccates were then resolubilised in 375 µl 10% D_2_O 1 mM non-deuterated DSS and recombined to make 1.5 ml solutions for NMR analysis.

The extracts were run on a 600 MHz Avance III NMR spectrometer (Bruker) with QCI-P cryoprobe at a calibrated temperature of 298K to acquire 1D-^1^H spectra. For each sample an automated program was set up on the spectrometer using ICON NMR including measurement of water offset, 90° pulse calibration, locking to D_2_O, tuning and shimming using an excitation sculpting experiment. A 1D-^1^H-NOESY was run with a mixing time of 100 ms, 512 scans and 8 dummy scans, a spectral width of 15.98 ppm (9.59 Hz), 32768 data points, an acquisition time of 2.27 s and a relaxation delay of 3 s.

The NMR spectra were phased, baseline corrected and had a 1 Hz exponential line broadening window function applied using TOPSPIN 3.6.1 (Bruker) software, then exported into Chenomx 8.4. The water resonance peak between 4.56 pmm and 4.97 ppm was deleted. The spectral peaks were then fit into the Chenomx library of metabolites using the profiler tool to match the peaks to their corresponding metabolites and concentrations.

Metabolites at significantly high abundances and of biological importance were divided into four groups; Short chain fatty acids (SCFAs), amino acids, sugars and sugar alcohols and other important metabolites and a time course was plotted to show the change in abundance of each metabolite throughout the antibiotic course.

## Results and Discussion

### Composition of gut bacterial communities and *Blastocystis* colonisation

Stool samples from days 0, 2, 3, 4, 5, 6, 7, 8, 10, 15, 30 days and 3 months after the start of the antibiotic course were collected and processed for both *Blastocystis* screening and 16S gut microbiome analysis. *Blastocystis* ST1 was present in samples from D2, D3 and consistently after D8 (**Table 1**).

**Table 1.** *Blastocystis* colonisation of stool samples was collected on different dates of the antibiotic course, 30 days, and three months after the completion of the course. + indicates the sample is *Blastocystis* + and – indicates the sample is *Blastocystis* –.

| Date of antibiotic course | *Blastocystis* +/- |
| --- | --- |
| Day 0 | - |
| Day 2 | + |
| Day 3 | + |
| Day 4 | - |
| Day 5 | - |
| Day 6 | - |
| Day 7 | - |
| Day 8 | + |
| Day 10 | + |
| Day 15 | + |
| 30 days after course | + |
| 3 months after course | + |

For the microbiome analysis, the most abundant genera (defined here as taxa whose mean abundance across all samples exceeded 1% of the total read count) across all stool samples were plotted as a heatmap and compositional plot (**Figures 1 – 2**). Seventeen genera met these criteria, while multiple genera showed patterns of change throughout the antibiotic course, including *Phocaeicola*_A, *Escherichia* and *Enterococcus*_B (**Figures 1 – 2**). Samples were also grouped by their similarity values (Euclidean distance scores) and ordered in a dendrogram. The bacterial communities of the samples collected 30 days and three months after the start of the antibiotic course were more similar in taxonomic distribution to the samples taken at both ends of the antibiotic course (D0-2 and D10-15) and had the least similarity to samples taken on D3-D7 (**Figure 1**). The samples collected on D0 and D2 were strongly similar to each other. *Bacteroides* and *Phocaeicola* (both from the phylum *Bacteroidota*) were the most abundant genera. They became more dominant throughout the first week of the course, with their relative abundances decreasing in the months after the completion of the course (**Figures 1** – **2**). *Phocaeicola* was the dominant genus and showed an increase in relative abundance in the first four days of the course, a slight decrease towards the end of the course and a further decrease in the months following the course (**Figure 2**). *Phocaeicola* was no longer the dominant genus after the antibiotic course, with *Escherichia* and *Enterococcus* being the most abundant in D30 and D3M, respectively (**Figure 2**). *Blastocystis* was not detected on D3, D4, D5, D6 and D7 of the course, but was again detected on D8 and recovered at the end. The presence of *Blastocystis* coincided with the decrease in abundance of *Bacteroides* and *Phocaeicola* (**Figure 2**). The reduced abundance of *Bacteroides* in the presence of *Blastocystis* is a consistent finding across studies globally [8], [14], [36], [37], [38], [39].

**Figure 1.**
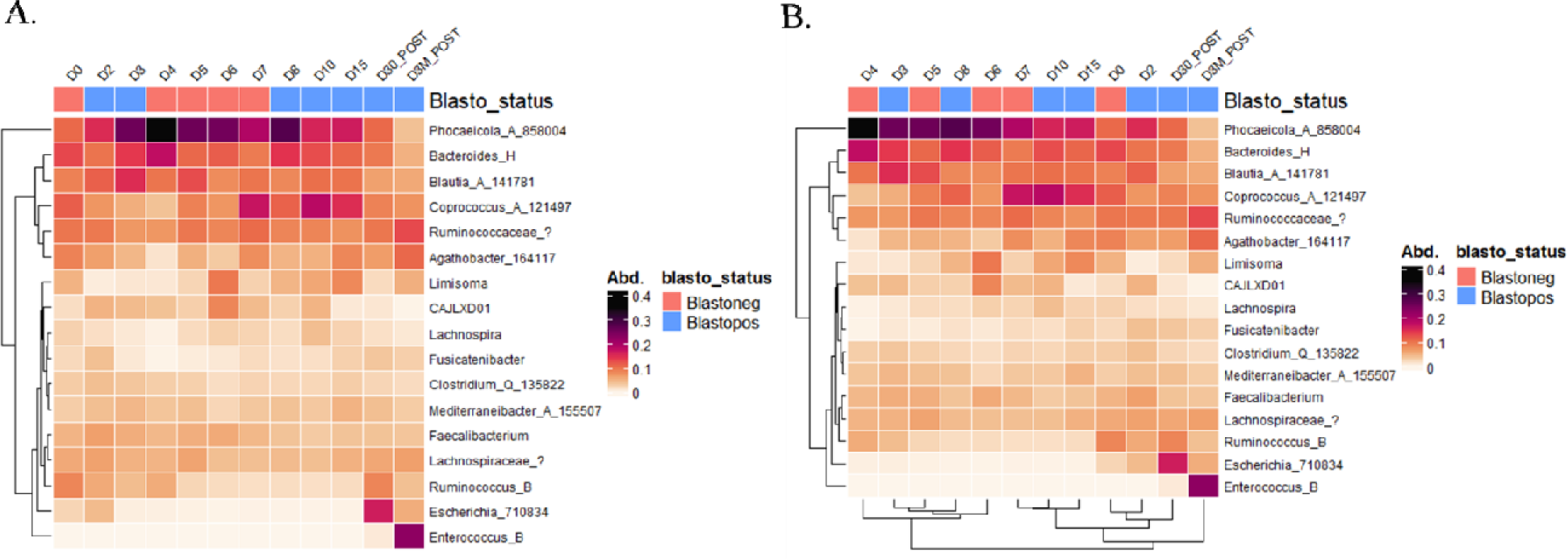
Heatmaps showing the relative abundance of taxa within the faecal samples, shown are all taxa which made up >1% of the total reads, all samples are indicated to be *Blastocystis* +ve (blue) or - ve (red) by a colour-coordinated legend. Dendrograms cluster both taxa (A-B) and the samples (B) using Euclidean distances and clustered using optimal leaf ordering. A). Heatmap showing the samples ordered by their collection timepoint during the antibiotics course. B). The heat map shows the samples ordered based on their similarity.

**Figure 2.**
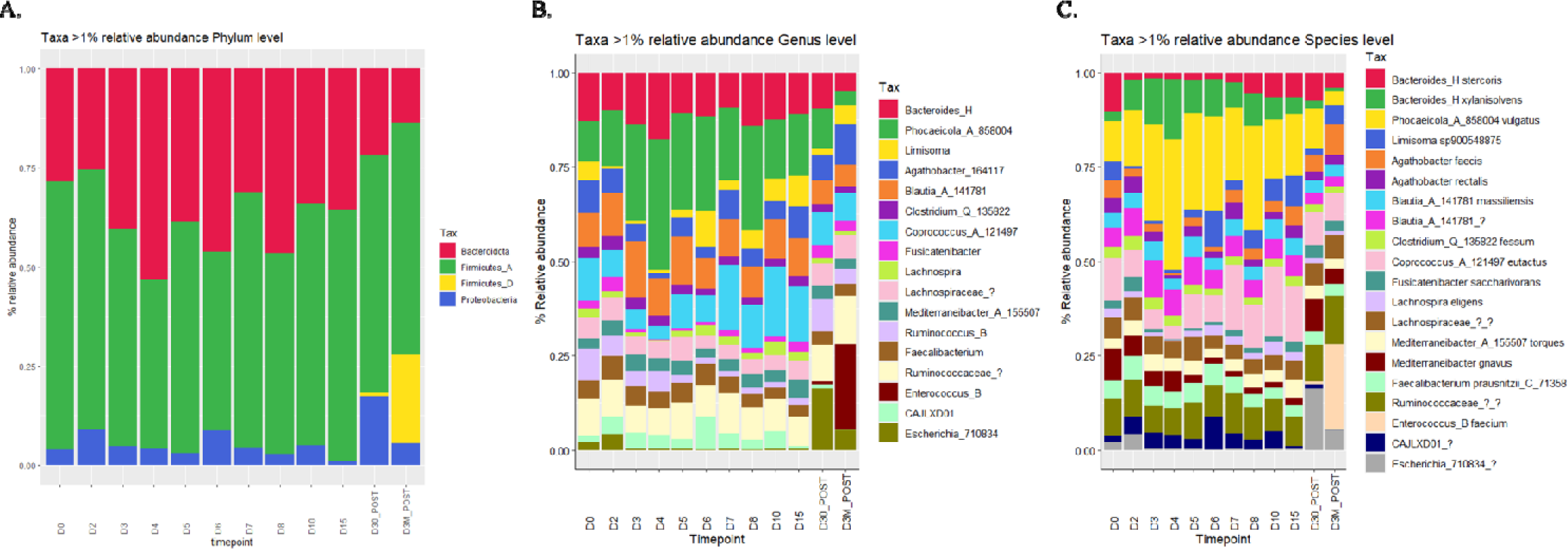
(A-C). Compositional plots showing the bacterial composition of the gut taxa aggregated to varying taxonomic levels. A). Phylum level B). Genus level C). species level. Taxa included made up >1% of total read counts respectively. Taxa abundances are shown as % relative abundance.

### Impact of antibiotic course on alpha diversity

To measure changes on alpha diversity, Shannon (factors in both evenness and richness), Chao1 (richness index which factors in potentially relevant singleton and doubleton ‘rare taxa’), Simpson (a measure of ‘dominance’/degree to which a few taxa make up most of the reads) and Observed taxa (true richness) indices were used. These diversity indices were analysed at the antibiotic-negative stage (pooled data of timepoints D0, D30, D3M) and at the antibiotic-positive stage (pooled data of timepoints D2, D3, D4, D5, D6, D7, D8, D10 and D15). All four metrics decreased during antibiotic administration. Antibiotics have been associated with acute gut microbiota perturbations defined by a decrease in taxonomic diversity [40], [41] (). However, these changes when analysed by ANOVA/Kruskal-Wallis were shown to be non-significant (>0.05 P-value) (**Supplementary Figure 1 B,D,F,H**).

The diversity metric scores were also plotted individually over time. The Shannon and Simpson diversity metrics decreased during the antibiotic course however the baseline composition recovered in the months after the course ended, although a decreasing trend was observed in D3M (**Figure 3 A, D**). This observation aligns with previous studies in both adults and children whereby core microbiome taxa return to their pre-antibiotics abundance [42]. Chao1 and Observed taxa showed sharp reductions at D4, D6 and D15. The reduction in these two indices on D15 contrasts the increase in the Shannon diversity score, indicating that the recovery in Shannon diversity score towards the end/post antibiotics course was characterised by a matching reduction in domination of the microbiome by a handful of taxa. This could be attributed to the elimination of rare taxa by the antibiotics.

**Figure 3.**
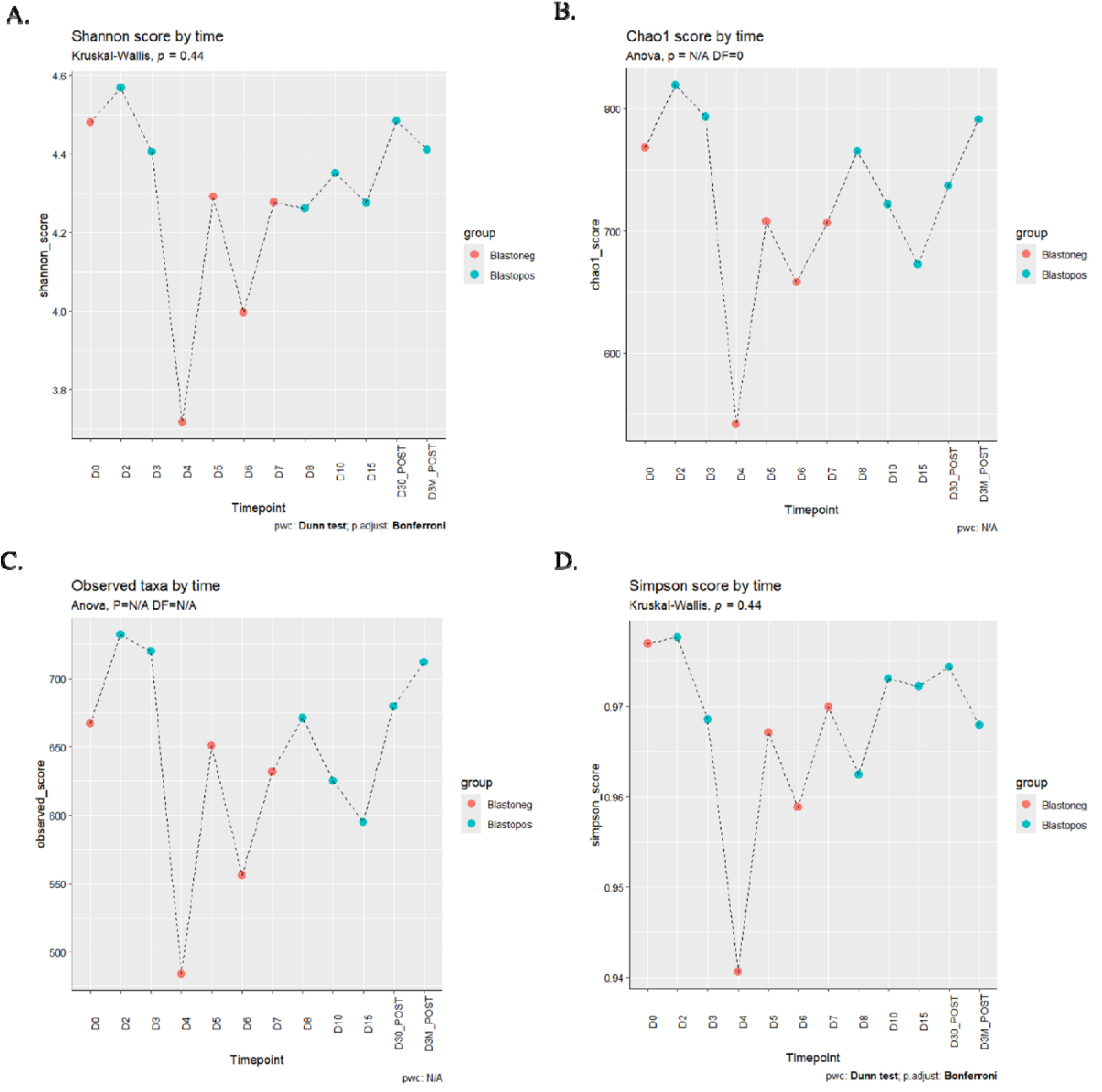
Statistical Diversity analysis of samples taken throughout the antibiotics course. A,B,C,D). Shannon, Chao1, Observed (richness), Simpson scores over time. Indicated if the sample was +ve (blue) or -ve (red) for *Blastocystis*.). Kruskal-Wallis H-test and Dunn’s test (Bonferroni p-adjust method) or ANOVA and Tukey-HSD test were used for statistical analysis (this was based on normality of the data, determined using the Shapiro test). Kruskal Wallis/ANOVA scores were all shown to be >0.05 indicating no-significance between the samples.

### Alpha diversity of Gut Microbial communities and *Blastocystis* colonisation

*Blastocystis* was detected early and later in the antibiotic course but not between D4 and D7 (**Figure 3**). All examined metrics of alpha diversity of *Blastocystis-*positive stool were increased when compared to the *Blastocystis-*negative stool (**Figure 3; Supplementary Figure 1**). These changes in diversity were analysed for statistical significance and found to all be non-significant (Observed taxa and Chao1 showed very low P-values of 0.058 and 0.064, respectively).

### Biomarker analysis of stool samples for *Blastocystis* colonisation

Having shown that (non-significant) increases in diversity occur with the presence of *Blastocystis* in stool samples, LEfSe was used to look for biomarkers of this change. LEfSE [35] locates taxa whose presence/absence allows for the best identification of a member of the Blastopos group instead of a member of the Blastoneg group [35]. Any LDA score >2 or <-2 is considered to be significant (**Figure 4**) shows all taxa with a significant LDA score and are coloured to show the indicative group. As shown in the plot, only two taxa were significantly viable at discriminating against the two groups in favour of *Blastocystis* negative samples (red, Blastoneg); these taxa were an unclassified member of the family *Anaerotignaceae* and the genus *Lactobacillus*. There was a considerably larger amount of taxa that were indicative of the *Blastocystis* positive group, with five taxa with a <-4 LDA score (**Supplementary Figure 2**); an unclassified member of the *Negativicutes* class of bacteria, an unclassified *Firmicute* bacteria, two different unclassified members of the family *Rikenellaceae* and an unclassified member of the family *Enterobacteriaceae*. The data suggests that some bacteria are indicators of the presence of *Blastocystis.* Still, the presence of so many indicative taxa may imply that a more diverse microbiome is the true indicative factor for *Blastocystis*.

**Figure 4.**
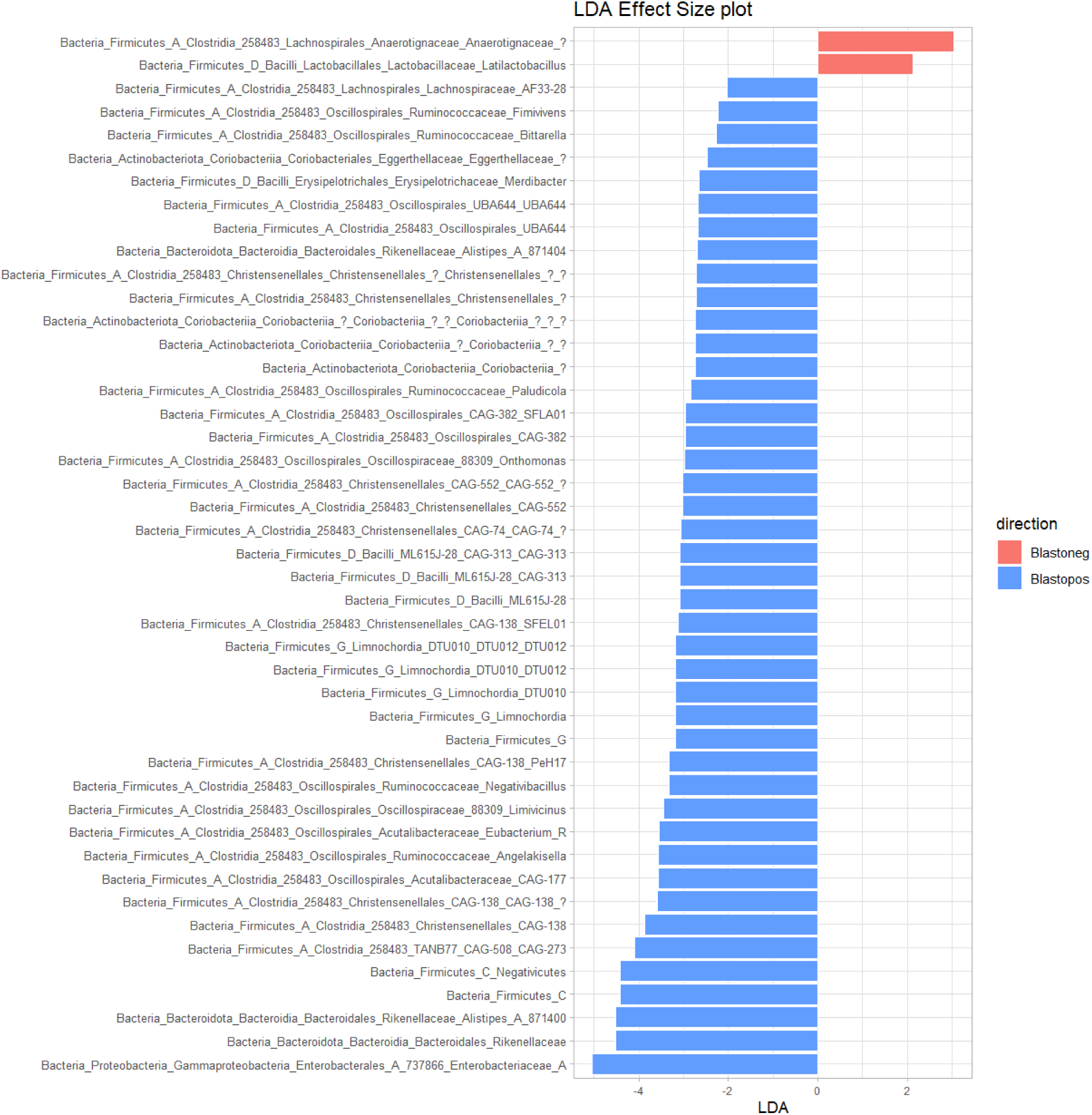
Linear Discriminant Analysis (LDA) Effect Size (LEfSe) plot. LDA scores indicate the presence/increased abundance of each taxa to discriminate between two conditions, *Blastocystis* +ve (blue) and *Blastocystis* -ve (red). Taxa with LDA scores between -2 and 2 are considered insignificant ‘biomarkers’ and are not included in the plot.

### Impact of antibiotic course on metabolite composition of the gut and *Blastocystis* **colonisation**

The metabolite extracts from the stool samples were subjected to 1D ^1^H NMR. Based on their chemical properties, four groups of metabolites were detected, including short-chain fatty acids (SCFAs) (**Figure 5a)**, amino acids (**Figure 5b)**, sugars and sugar alcohols (**Figure 5c)** and others (**Figure 5d**). Previous metabolome investigations at a single timepoint on *Blastocystis* positive and negative individuals showed a decreased abundance of certain metabolites in the former [2]. Specifically, Alanine, Glycine, Histidine, Isoleucine, Methionine, Threonine, Tryptophan and Valine all decreased, suggesting an anti-inflammatory role of *Blastocystis*. Significant increases in certain amino acids in the stool have been found in IBD patients [43]. Herein, in a time course metabolome of a single individual, all the amino acids, particularly Alanine and Valine, showed a decrease mid-course and recovery towards the end of the course.

**Figure 5.**
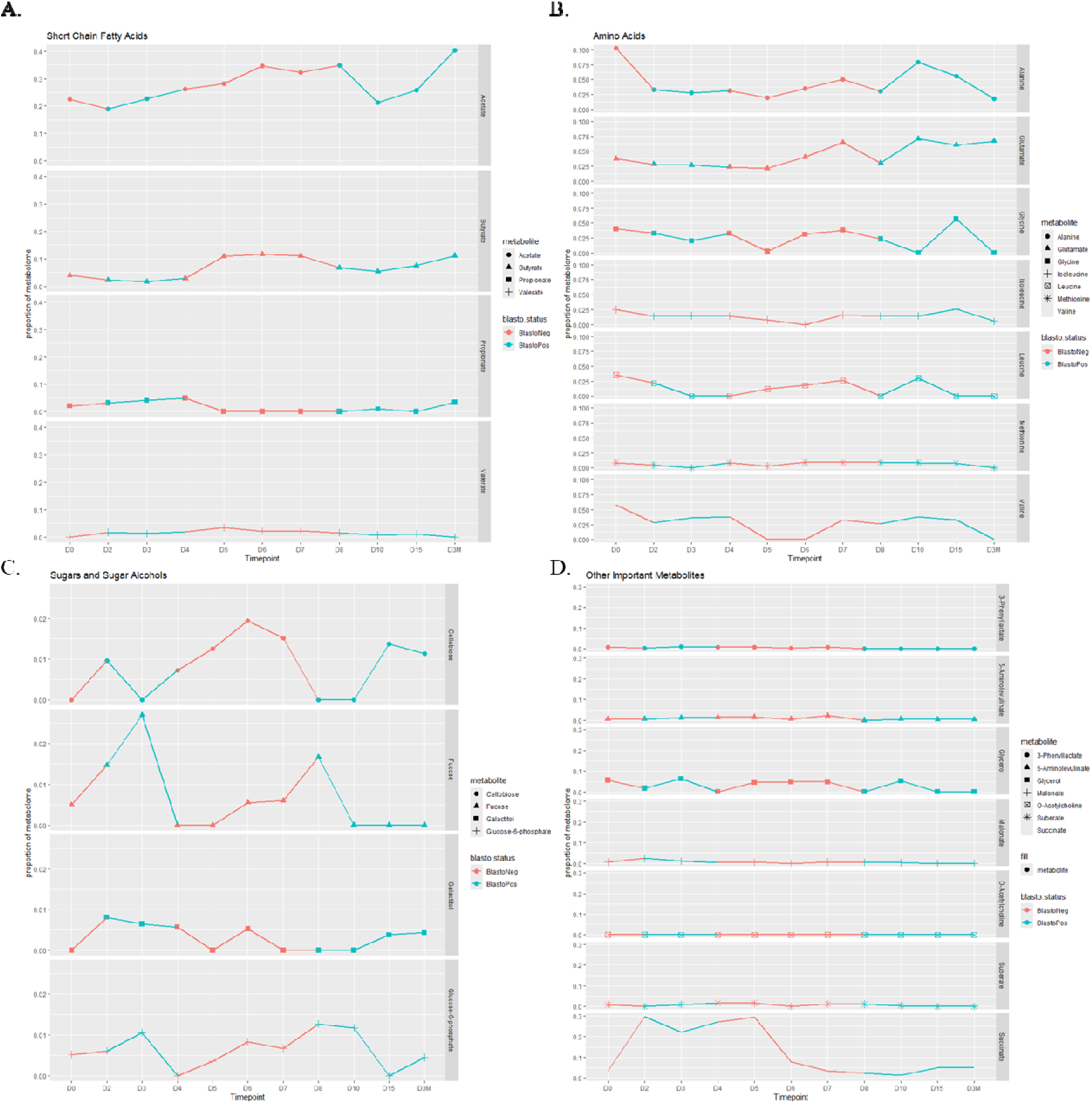
Time course of metabolite abundances of four different groups of metabolites throughout the antibiotic course as well as *Blastocystis* Colonisation. a. SCFAs b. Amino acids. c. Sugars and Sugar alcohols d. Other important metabolites

Moreover, all amino acids, but glutamate showed a large decrease post-course (**Figure 5b**). Regarding SCFAs, the abundance of acetate increased throughout the first week of the antibiotic course but decreased during the second week and then recovered post-course. Butyrate increased after the first four days, decreased on D7, and recovered post-course. Whether these alterations reflect changes in absorption or loss remains an open question. Cellobiose was the most impacted sugar by the antibiotic course and showed a large increase in abundance during the first week, then decline and recovery during the second week (**Figure 5c**). Malonate steadily declined for the first five days and was undetectable by D6. It then recovered on D7, D8 and D10 but became undetectable on D15 and post-course (**Figure 5d**). Succinate sharply increased from D1 to D2 and stayed high until D5, when it declined again. O-acetylcholine declined for the first five days then recovered on D6 and D7, but disappeared during the second week and didn’t recover post-course. Notably, acetylcholine in the gut plays a role in intestinal homeostasis; hence, its disruption could potentially aggravate inflammation [44].

### Conclusion

This case study provides valuable insights into the impact of antibiotic treatment on the presence of *Blastocystis* and the overall gut microbiome and metabolome. The 14-day course of Lansoprazole, Amoxicillin, and Clarithromycin significantly altered the gut microbial composition, causing a notable decline in microbial diversity mid-course and leading to a temporary absence of *Blastocystis*. Despite the antibiotic-induced perturbations, *Blastocystis* demonstrated resilience, re-emerging post-treatment.

Additionally, the study highlighted significant fluctuations in metabolite levels, including short-chain fatty acids and amino acids, which are critical for gut health. These changes underscore the profound influence antibiotics have not only on microbial populations but also on the metabolic environment of the gut. While antibiotics did not have a lasting effect on *Blastocystis* colonisation, their temporary impact on microbial diversity and metabolite composition points to gut ecology’s intricate and dynamic nature. The findings emphasise the need for careful consideration of antibiotic use, especially in conditions like IBS, where maintaining a balanced gut microbiome is crucial.

Future research should expand on these findings by exploring the long-term effects of antibiotics on gut microbiota and metabolites in larger cohorts. Understanding these interactions will be essential for developing targeted therapies that mitigate adverse impacts on the gut ecosystem while effectively treating gastrointestinal conditions.

## Data Availability

All data produced in the present study are available upon reasonable request to the authors

## Acknowledgements

We would like to thank the volunteer for participating in this study. Many thanks to the Tsaousis Lab members (2020 – 2021) for their help and support in sample collection and managing this project. J.M.N. was supported by a Kent Health studentship and W.J.S.E. by a SoCoBio DTP studentship.

## Contributions

Conceptualisation, A.D.T.; methodology, J.M.N., W.J.S.E. and G.S.T.; software, G.S.T.; validation, G.S.T. and W.G. and W.J.S.E..; formal analysis, J.M.N. and W.J.S.E.; investigation, J.M.N. and W.J.S.E.; resources, A.D.T.; data curation, J.M.N. and E.G.; writing—original draft preparation, J.M.N.; writing—review and editing, A.D.T., E.G.; supervision, A.D.T.; project administration, A.D.T.; funding acquisition, A.D.T. All authors have read and agreed to the published version of the manuscript.

## Notes

### Competing Interest Statement

The authors have declared no competing interest.

### Funding Statement

This study did not receive any funding

### Author Declarations

The National Ethics committee [Health Research Authority (HRA) and Health and Care Research Wales (HCRW)] of the NHS Health Research Authority at the United Kingdom gave ethical approval for this work.

