## Supplementary Figure for "The impact of antibiotics on the presence of the protozoan anaerobe *Blastocystis* and the surrounding microbiome: a case study"

### Supplementary figures

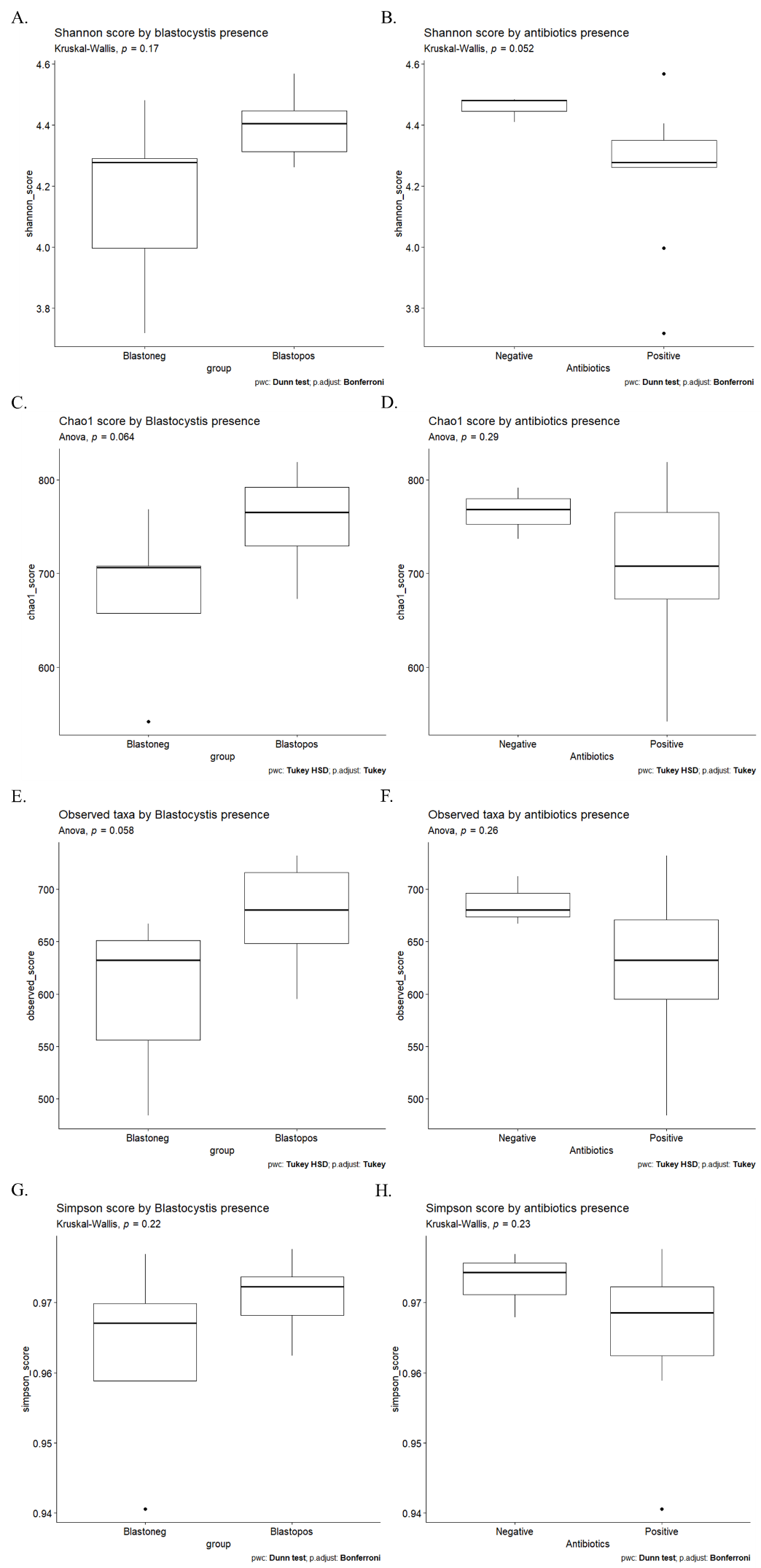

**Supplementary figure 1.** A, C, E, G). Statistical analysis comparing samples that were +ve or -ve for *Blastocystis*. B, D, F , H). Statistical analysis comparing samples taken during the course of antibiotics and samples taken outside of the course (D0 before, 30 days after and 3 months after). Kruskal-Wallis H-test and Dunn’s test (Bonferroni p-adjust method) or ANOVA and Tukey-HSD test were used for statistical analysis (this was based on normality of the data, determined using the Shapiro test). Kruskal Wallis/ANOVA scores were all shown to be >0.05 indicating no-significance between the samples.

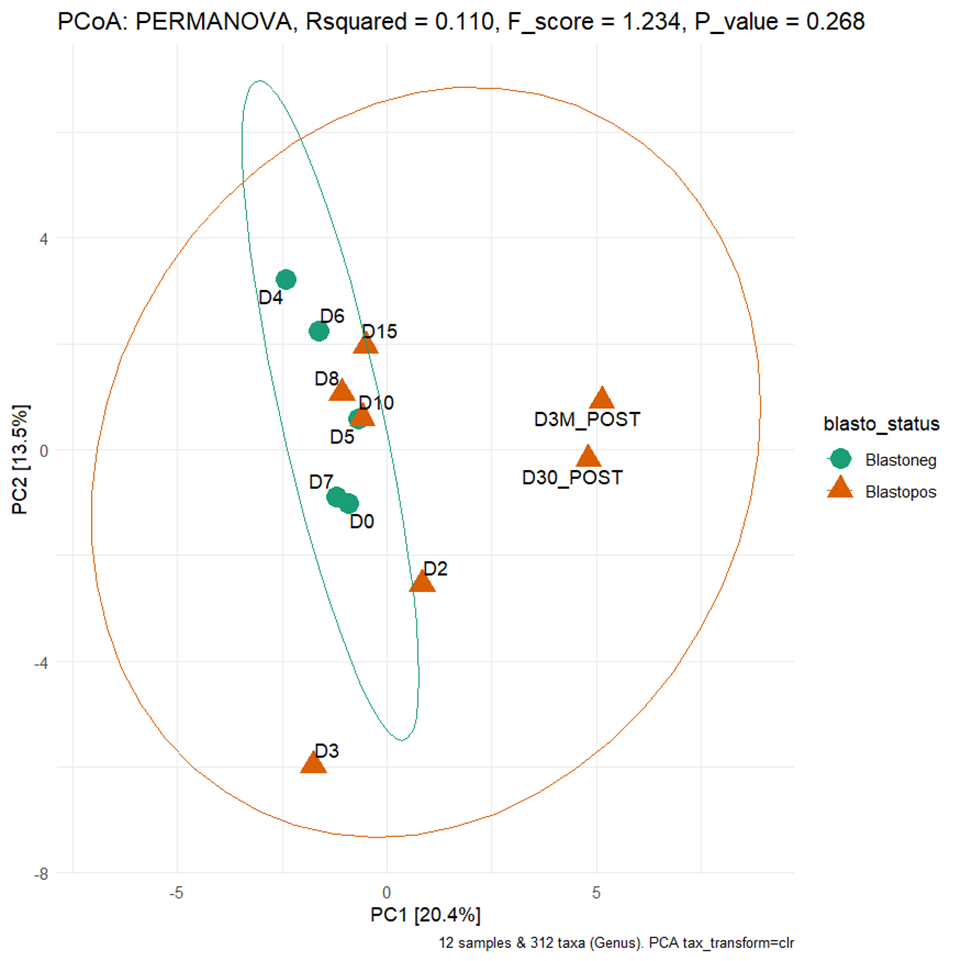

**Supplementary Figure 2.** Principle component analysis (PCA) plot. The plot shows the positions of each sample in a dissimilarity matrix (Euclidean distances). Different groups are indicated by shape and colour: *Blastocystis* -ve (circle, green) and *Blastocystis* +ve (triangle, red). Statistical analysis of the different groups’ positions was performed using PERMANOVA. The PERMANOVA P-value >0.05 indicates there is no significant difference between the positions of each group's centroid.
